# Safety and efficacy of rapamycin on healthspan metrics after one year: PEARL Trial Results

**DOI:** 10.1101/2024.08.21.24312372

**Authors:** Girish Harinath, Virginia Lee, Andy Nyquist, Mauricio Moel, Stefanie L. Morgan, Anar Isman, Sajad Zalzala

## Abstract

**Background:** Low-dose rapamycin promotes longevity in mice, but clinical safety and longevity data effects in humans remain limited.

**Objectives:** Evaluate the long-term safety of intermittent low-dose rapamycin in a healthy, normative-aging human cohort.

**Design:** This decentralized double-blinded, randomized, placebo-controlled trial (NCT04488601, registered 2020-07-28) was performed over 48 weeks. Participants received placebo, 5mg or 10mg compounded rapamycin (equivalent to 1.43mg or 2.86mg of generic formulations) weekly. The primary outcome measure was visceral adiposity (by DXA scan), secondary outcomes were blood biomarkers, and lean tissue and bone mineral content (by DXA scan). Established surveys were utilized to evaluate health and well-being. Safety was assessed through adverse events and blood biomarker monitoring.

**Results:** Adverse and serious adverse events were similar across all groups. Visceral adiposity did not change significantly (η_p_^2^=0.001, *p*=0.942), and changes in blood biomarkers remained within normal ranges. Lean tissue mass (ε^2^=0.202, *p*=0.013) and self-reported pain (ε^2^=0.168, *p*=0.015) improved significantly for women using 10mg rapamycin. Trends of improvement in bone mineral density were observed in males using 10mg rapamycin (ε^2^=0.221, *p*=0.061), but no other significant effects were observed.

**Conclusions:** Low-dose, intermittent rapamycin administration over 48 weeks is relatively safe in healthy, normative-aging adults, and was associated with significant improvements in lean tissue mass and pain in women. Future work will evaluate benefits of a broader range of rapamycin doses on healthspan metrics for longevity, and will aim to more comprehensively establish efficacy.

## Introduction

Aging is the greatest risk factor for all major chronic diseases, accounting for nearly 70% of human mortality [1–3]. While advancements in medical technologies and public health practices over the past 150 years have led to longer lifespans less shaped by natural selection, the period of disease and disability-free life often referred to as “healthspan” has not kept pace [4]. In conjunction with an epidemic of poor lifestyle habits, this has collectively led to a growing chasm between lifespan and healthspan known as the healthspan gap, which in the United States lasts several decades and is characterized by a high burden of functional disability and age-related diseases (such as type 2 diabetes, osteoarthritis, and Alzheimer’s) that often coexist as multi-morbidities [5]. While significant research has historically focused on treating these diseases individually, a growing body of work within translational geroscience explores developing gerotherapeutics that slow the aging process and delay the onset of or prevent age-related disease altogether[6].

The field of translational geroscience has made rapid advancements in recent years, due in large part to the strategic utilization of interventions already approved for other conditions by the US Food and Drug Administration (FDA) [7]. By repurposing such drugs for their potential to target the biology of aging and extend healthy longevity, clinical validation is fast-tracked to permit a more immediate collection of application-specific efficacy data. Notable among these is rapamycin, which is widely used for its purported longevity and healthspan benefits within the pro-longevity community [8]. While evidence supports a role for rapamycin in improving life- and health-spans in preclinical studies [9], little data exists on its clinical efficacy in normative aging humans.

As an FDA-approved small molecule drug, rapamycin is an evolutionarily conserved inhibitor of the mammalian target of rapamycin serine/threonine kinase complex 1 (mTORC1), though it is known to also impact mTORC2 in certain contexts. mTORC1 is a known regulator of aging processes, and its hyperactivity has been linked to multiple chronic disease processes [10, 11]. Conversely, partial inhibition of mTORC1 induced by caloric restriction and rapamycin is hypothesized to be a major mediator of their lifespan and healthspan-enhancing effects across organisms from yeast to non-human primates [12–21]. Rapamycin has demonstrated particular efficacy as a geroprotective intervention in mice, extending lifespan in heterogeneous genetic backgrounds across multiple studies from independent labs at multiple dosages, dosing periods, and regimens, even in elderly animals [14, 16, 21, 22].

While rapamycin has demonstrated a more pronounced lifespan-enhancing effect in female than male mice, mitigation or reversal of age-associated changes in multiple organ tissues and the immune system have been observed for both sexes [23–25]. Preliminary data suggests rapamycin effects on mitigating age-related decline are conserved in companion dogs and marmosets, however, clinical data on rapamycin’s gerotherapeutic effects in humans remains limited [9, 12, 17, 26]. Given the substantial promise in preclinical data, it is essential to obtain a deeper understanding and clearly define the clinical benefits of rapamycin use for improving healthy aging in the generally healthy, adult population. In particular, it will be important to understand how rapamycin may impact the phenotypes of the biological aging process that substantially increase the risk of age-related disease and mortality, such as an accumulation of visceral adipose tissue (VAT) and a loss of lean muscle tissue and bone mass [27–32]. These are some of the most salient and functionally consequential measures of biological aging, which frequently lead to reduced quality of life (QoL), increased pain, and limited mobility, particularly for post-menopausal women [33, 34]. Collectively, this contributes to the marked decline in health and increased incidence of age-related disease that often occurs during this time period. It has been suggested that use of low-dose rapamycin may mitigate these features of the aging process, enhancing healthspan [25, 35, 36].

The widespread adoption of rapamycin as a gerotherapeutic has historically been limited by concerns regarding its known impact on immunosuppression, hyperlipidemia, and hyperglycemia [37]. However, the vast majority of these effects stem from chronic daily dosing regimens utilized in severely ill organ transplant or cancer patients, where the clinical aim is inhibition of the immune system or anti-tumorigenic effects. In contrast, as a gerotherapeutic for normative aging populations, low-dose, intermittent rapamycin is revealing promise for minimizing side effects while still mitigating aspects of age-related decline [9, 38, 39]. For example, Mannick et al. demonstrated that healthy elderly individuals taking 0.5 mg of a rapalog daily or 5 mg/week for 6 weeks mitigated age-related immune decline by enhancing the adaptive immune system’s response to vaccination [39]. This supports our recent findings from a study of 333 low-dose rapamycin users indicating a high perceived QoL and improved health outcomes compared to non-users [8]. Preliminary data suggest that these effects may be driven by partial inhibition of mTORC1 with low-dose, intermittent rapamycin (rather than the complete inhibition observed with chronic dosing regimens), which may mitigate age-related decline by stimulating autophagy, reducing senescent burden, and reducing pro-inflammatory cytokines that drive chronic inflammation [9, 15, 24, 40, 41]. While promising findings such as these have encouraged some physicians to prescribe off-label rapamycin as a therapy to maintain healthspan, there are many open questions that require further study, particularly in a clinical setting.

An important gap in the clinical understanding of rapamycin for longevity is that to date, no long-term randomized controlled trials (RCT) have been conducted to explore low-dose, intermittent rapamycin regimens for improving multiple healthspan metrics in normative aging cohorts. The current study, the Participatory Evaluation of Aging with Rapamycin for Longevity (PEARL) trial, aimed to address this gap. PEARL represents the longest clinical study of rapamycin use for healthy aging performed to date. This 48-week double-blinded, randomized placebo-controlled decentralized study evaluated the safety and efficacy of 5 and 10 mg/week compounded rapamycin in mitigating clinical signs of aging in a normative aging cohort. Importantly, we determined in a previous study that these dosages are equivalent to 1.4 and 2.9 mg/week of commercially available rapamycin, representing a very low dosage schema compared to previously published literature. Here, we present the first evidence to date, to our knowledge, that low-dose rapamycin administration is safe over the course of 48 weeks. Further, while our primary endpoint of visceral adiposity showed no significant changes over the course of the study, secondary endpoints of lean muscle mass and self-reported pain significantly improved for women in the 10mg rapamycin cohort. Despite small participant numbers, these effects combined with the relatively low doses utilized here provide sufficient evidence that further investigation into rapamycin as a beneficial gerotherapeutic are warranted.

## Methods

### Study design

The PEARL study was a decentralized, single-center, prospective, double-blind, placebo-controlled trial assessing rapamycin in healthy individuals aged 50-85 years, to determine the safety and efficacy in mitigating aging-related decline (**Supplementary Figure S1**). It was registered as a clinical trial on 2020-07-28, NCT04488601, and was conducted in accordance with the standards of Good Clinical Practice, as defined by the International Conference on Harmonisation and all applicable federal and local regulations. The study protocol was approved by the institutional review board of the Institute of Regenerative and Cellular Medicine in May 2020 (IRCM; approval number IRCM-2020-252). Discussions with the FDA determined that this study was exempt from IND requirements.

### Study endpoints

The primary endpoint of this study was changes in visceral fat as measured by dual-energy x-ray absorptiometry (DXA) scan. Secondary endpoints included changes in lean tissue mass and bone density as determined by DXA scan, as well as changes in blood biomarkers from complete blood count (CBC), blood electrolytes, liver function, renal function, serum glucose, insulin, and hemoglobin A1c. Standardized self-reported surveys of quality of life (SF36, [42]) and frailty (WOMAC, [43]) were also completed by study participants, but were not included as specific study endpoints.

### Study population

Participants were recruited and screened for eligibility via the AgelessRx online medical platform. If deemed eligible, informed consent was obtained for participation in the study. Participants were eligible for the study if they were aged between 50 and 85 years at the start of the study, were interested in taking rapamycin off-label, were willing to undergo minimally invasive tests, and were in good health or had well-managed clinically-stable chronic diseases. Participants were excluded from the study if they had anemia, neutropenia, or thrombocytopenia, were premenopausal, were scheduled to undergo major surgery in next 12 months, were undergoing or were scheduled to undergo chemotherapy, were scheduled for immunosuppressant therapy for an organ transplant, had impaired wound healing or history of chronic open wound, untreated dyslipidemia, impaired hepatic function, chronic infections requiring ongoing treatment or monitoring (e.g. human immunodeficiency virus/acquired immunodeficiency syndrome, chronic Lyme disease), allergy to rapamycin, clinically-relevant primary or secondary immune dysfunction or deficiency, chronic oral corticosteroid or immunosuppressive medication use, fibromyalgia, chronic fatigue syndrome/myalgic encephalitis, breast implant illness, congestive heart failure, impaired renal function, poorly controlled diabetes, type I or insulin-dependent type II diabetes, untreated or treated within the last five years for substance abuse disorder, and untreated or poorly controlled mental health disorder. Further, those who had recently taken or were taking metformin, rapamycin, or rapalogs were excluded unless the participant agreed to a 6 month washout period prior to the start of the trial.

### Treatments

Rapamycin used in this study was a compounded formulation of 5mg or 10mg, received from Belmar Pharma Solutions (Golden, CO, USA). Placebo capsules were also formulated by Belmar, and were designed to have a similar appearance to rapamycin capsules. Both were taken orally. Importantly, data received since this study was completed suggest that this particular compounded rapamycin formulation is approximately 3.5x less bioavailable than the standard generic rapamycin (also known as Sirolimus, or by the brand name Rapamune). As such, the doses used in this study were likely approximately equivalent to 1.43mg and 2.86mg of the generic product. For simplicity, we refer to doses in this manuscript at their compounded levels of 5mg and 10mg.

Study participants were randomized into three groups: receiving 5 mg of compounded rapamycin, 10 mg of compounded rapamycin, or placebo, once per week by mouth. Participants were instructed they could take the medication with or without food. Study participants were prescribed the drug for 48 weeks upon enrollment in the study and were dispensed supplies for 12 weeks at a time. After completion and unblinding of the trial, participants receiving the placebo were given 1 year of no-cost compounded rapamycin if desired, and were monitored for any adverse side effects.

### Assessments

All assessments were performed at baseline, after 24 weeks, and after 48 weeks of rapamycin treatment, and included comprehensive blood testing, DXA body composition scans, and established self-report surveys (SF36 and WOMAC scales). Safety-based blood testing (including Triglycerides, Total Cholesterol, LDL-Cholesterol, Glucose, Creatinine, ALT, WBC, RBC, Hemoglobin, Hemoglobin A1C, and ApoB) was performed two additional times (at 2 weeks and 4 weeks of treatment for all markers except Hemoglobin A1C and ApoB, which were limited to baseline, 24 weeks, and 48 weeks) to evaluate safety. All blood testing was performed by local Quest Diagnostics or LabCorp laboratories, and included complete blood count (CBC), comprehensive metabolic panel, liver function tests, renal function tests, lipid panels, and insulin/glucose monitoring panels. Participants were asked to fast the night before blood draws.

DXA scans were performed by designated partner facilities DexaFit and Fitnescity at locations convenient for the participants, and were used to measure visceral adiposity, bone density (from both bone mineral content and bone mineral density), and lean tissue mass. AgelessRx staff assisted participants in finding and scheduling appointments at these facilities as needed. For participants for whom neither partner facility had a convenient nearby location, alternative facilities were identified in conjunction with the AgelessRx staff. For all DXA scan facilities, scans were completed by trained technicians familiar with equipment calibration and function, appropriate patient positioning, and necessary safety protocols. For all participants, the following procedures were used:

Pre-Scan, participants were advised to avoid calcium supplements and certain medications for 24-48 hours before the scan to prevent interference with results. They were asked to avoid exercise prior to the scan and come in well-hydrated. Additionally, they are to fast 2-3 hours prior to their scan, and inform the technician of any recent surgeries, fractures, or medical conditions that may impact the scan. During the scan, they were asked to wear loose, metal-free clothing, and to remove metal objects such as jewelry or belts to avoid artifacts.

All DXA measures are obtained by comparing the X-ray attenuation in the designated region or tissue type. Measurements are reported in grams or grams per cm^2^. Results are compared to those of national averages for a participant’s given age, gender, and race.

Measures of gut microbiome health were evaluated using the at-home Thorne Gut Health test (Thorne), and measures of epigenetic age were evaluated using the at-home TruDiagnostic TruAge kit (TruDiagnostic). Results were provided with the kit, and interpreted versions were returned to AgelessRx researchers for correlation and comparison with dosing groups and other measures reported in this manuscript.

Health-related QoL was assessed by the short-form 36 (SF-36) survey, which consists of 36 questions covering eight health domains: physical functioning, role limitations due to physical health, bodily pain, general health perceptions, vitality (energy levels), social functioning, role limitations due to emotional problems, and emotional health [42]. The responses are scored and summarized to provide a profile of an individual’s perceived health status. Pain, fitness, and functional limitations were assessed using the Western Ontario and McMaster Universities Osteoarthritis (WOMAC) index, which is a questionnaire that is commonly used to assess the health status of individuals with osteoarthritis of the hip and knee [43]. It consists of 24 items divided into three subscales: pain, stiffness, and physical function. The responses are scored to provide quantitative assessments of the severity of symptoms and functional limitations associated with osteoarthritis.

### Participant Protocol Adherence and Adverse Event Monitoring

Participants were monitored routinely throughout the study through weekly email surveys and 4 virtual meetings. They were asked to schedule all 4x virtual check-in meetings (via Calendly) with clinical trial staff at the time of onboarding. Timestamps for the meetings include 2 weeks (after 1st dose date), 4 weeks, 6 months, and final 12-months. During these meetings, any outstanding items that participants had not yet completed would be reviewed and documented. Items included verifying weekly administration of their medication, completing weekly check-in surveys (WOMAC, SF36, adverse events, etc), completing at-home kits (as relevant), scheduling and completing Quest blood draws as required, and completing DXA scans as required. Clinical trial staff had internal project management sheets to monitor participants and their scheduled tasks, as they progressed through trial. Any missed meetings due to participant schedule conflicts/no-shows/cancellations would be re-scheduled or followed up in a timely manner with participants to ensure trial compliance and support as needed.

Adverse events (AEs) were obtained through weekly monitoring forms sent out to participants. Clinical trial staff reviewed and documented all AEs, and conferred with medical staff as necessary to determine if individuals should be removed from the study for any specific AEs or serious AEs. A full list of AEs, withdrawn patients, and SAEs is presented in **Supplementary Table S1**.

### Statistical analyses

Data were analyzed using tests as described in the text, with relevant corrections for sphericity (Greenhouse-Geisser), homogeneity of variance (Welch’s test), and multiple comparisons (Bonferroni or Games-Howell correction), unless otherwise noted. All analyses were conducted using SPSS 29.0.2.0 (IBM, Armonk, NY, USA). Not all participants completed all datapoints, thus in some tests, the number of cases will differ from the total number of study participants. For each test, the maximum number of datapoints was utilized for comparisons, with pairwise removal for missing values.

## Results

A total of 114 participants completed the study and were included in data analysis. An additional 11 discontinued participation prior to study completion, and were not included in these analyses. Of the 114 who completed the study, 40 received 5 mg/week of rapamycin, 35 received 10 mg/week of rapamycin, and 39 received placebo (**Supplementary Fig S1)**. Participant dosing groups were not significantly different at baseline on the vast majority of measures, including age, gender, weight, and BMI, however, we observed a relatively low enrollment of women across all groups (35.1% of participants overall (n = 40), with 20% in the 10mg group (n = 8), 42.5% in the 5mg group (n = 17), and 38.5% in the placebo group (n = 15; **Supplementary Table S2**). Additionally, we noted that at baseline, ANOVA (with Bonferroni corrected post-hoc testing, unless otherwise noted) suggested that participants in the 10mg group had significantly higher scores on SF-36 self-report survey measures of both emotional well-being (*F(2, 110) =* 4.083, *p* = 0.019, *ε^2^*= 0.052; *md* = 7.667 (95% CI = 0.970 - 14.364), *p* = 0.019) and role limitations due to emotional problems (Welch’s ANOVA *F(2, 60.793) =* 4.103, *p* = 0.021, *ω^2^* = 0.065; Games-Howell *md* = 10.653 (95% CI = 0.706 - 20.599), *p* = 0.033; **Supplementary Table S3**). Similarly, the 5mg group had a slightly lower hemoglobin A1C than placebo at baseline (by ANOVA with Bonferroni corrected post-hoc tests, *F(2, 106) =* 3.418, *p* = 0.036, *ε^2^* = −0.015; *md* = −0.1497 (95% CI = −0.289 - −0.010), *p* = 0.031), despite self-reporting less moderate activity (with Welch’s ANOVA and Games-Howell post hoc tests, *F(2, 65.313) =* 5.315, *p* = 0.007, *ω^2^* = 0.076; *md* = −0.568 (95% CI = −1.07 - −0.070), *p* = 0.023), and had a significantly lower baseline measure of bone mineral density by DXA scan (*F(2, 70.147) =* 4.250, *p* = 0.018, *ω^2^* = 0.104) relative to both 10mg (*md* = −0.19684 (95% CI = −0.3492 - 0.0345), *p* = 0.014) and placebo groups (*md* = −0.16915 (95% CI = −0.3333 - −0.0049), *p* = 0.042; **Supplementary Table S3**).

For those who discontinued participation, 6 were in the placebo group, 3 in the 10mg group, and 2 in the 5mg group. Comprehensive details regarding participants who optionally withdrew or who experienced serious adverse events (SAEs) are included in **Supplementary File 1** and **Supplementary Table 1**, and include 1 event in the 10mg group, 2 in the 5mg group, and 3 in the placebo, of which only one placebo user was withdrawn (**Supplementary Figure S2a**). For non-severe adverse events (AEs), similar total numbers were reported in all groups (10mg = 117, 5mg = 116, placebo = 122), with no clear differences by gender (10mg: Female = 48, Male = 69, 5mg: Female = 57, Male = 59, placebo: Female = 76, Male = 46; χ^2^ of all comparisons non-significant). As some participants reported multiple AEs, we compared the number of participants reporting AEs (**Supplementary Figure 2b**). This was also found to be relatively consistent across all groups (10mg = 29 (80.6%), 5mg = 31 (77.5%), and placebo = 34 (87.2%)) and genders (10mg: Female = 8 (88.9%), Male = 21 (77.8%); 5mg: Female = 13 (76.5%), Male = 18 (78.3%); placebo: Female = 13 (86.7%), Male = 21 (87.5%); **Supplementary Figure S2c**). AEs were relatively consistent across all groups, though GI symptoms were reported more often for rapamycin users than placebo (10mg = 8, 5mg = 7, placebo = 4).

Phenotypic hallmarks of biological aging were evaluated using DXA scans of body composition after 24 and 48 weeks of treatment, specifically for measures of visceral adipose tissue (VAT), bone mineral content (BMC), bone mineral density (BMD), and lean tissue mass (LTM). Given expected differences in participant body composition and size at baseline (i.e., participants spanned a 43.18 cm range in height, 74.2 kg in weight, and BMI from 18.5 – 36.5; **Supplementary Table S2**), all DXA-based body composition measures were normalized to individual baseline as a percent change over the described time before further analysis.

Following this normalization, odds ratios were calculated for all composition measures, specifically for the primary endpoint of visceral fat changes as determined by DXA visceral adipose tissue measurements. As outlined in **Table 1**, odds ratios for visceral fat suggested both 10mg (*OR* = 1.43, 95% CI = 0.48 - 4.25) and 5mg (*OR* = 1.95, 95% CI = 0.66 - 5.79) rapamycin users were more likely to see improvements in measures of visceral fat than placebo users, with benefits for females in both groups (10mg: *OR* = 5.63, 95% CI = 0.50 - 63.28; 5mg: *OR* = 4.41, 95% CI = 0.39 - 49.63), but males only in the 5mg group (10mg: *OR* = 0.89, 95% CI = 0.25 - 3.13; 5mg: *OR* = 1.57, 95% CI = 0.44 - 5.51). Similar trends were seen for secondary outcome measures of lean tissue mass, with all rapamycin users in all groups more likely to have benefits than placebo (see **Table 1**). In contrast, secondary outcome measures of bone density, as evaluated by DXA bone mineral density (BMD) and bone mineral content (BMC) measures, suggested that while 10mg rapamycin users saw more improvements than 5mg rapamycin users on BMD, this effect was specific to men, and for BMC, rapamycin users were more likely to lose bone density overall compared to placebo users (see **Table 1**). Despite these suggestions from ORs, repeated measures mixed ANOVA of DXA-based body composition changes at 24 and 48 weeks by dosing group showed no significant change for dosing group over time for any measure (VAT: *F(2, 85)* = 0.060, *p* = 0.942, LTM: *F(2, 85)* = 0.751, *p* = 0.475, BMC: *F(2, 82) =* 0.470, *p* = 0.627, BMD: *F(2, 107)* = 0.573, *p* = 0.565; **Supplementary Table S4**). Findings of non-significance with this approach were not changed after simplifying the analytical approach (likely due to the power limitations inherent to this cohort and significant variability in individual response on each measure across all groups) by evaluating changes within each group across time with repeated measures ANOVA, or when separating by gender with either analysis method (**Supplementary Table S4, Supplementary Figure S3a-h**).

**Table 1.** Odds Ratios of Improvement on Body Composition Metrics.

| Table 1. Odds Ratios of Improvement on Body Composition Metrics |  |  |  |  |  |  |
| --- | --- | --- | --- | --- | --- | --- |
|  | Odds Ratios |  |  |  |  |  |
|  | All |  | Female |  | Male |  |
| VAT | OR | 95% CI | OR | 95% CI | OR | 95% CI |
| 10mg | 1.43 | 0.48 - 4.25 | 5.63 | 0.50 - 63.28 | 0.89 | 0.25 - 3.13 |
| 5mg | 1.95 | 0.66 - 5.79 | 4.41 | 0.39 - 49.63 | 1.57 | 0.44 - 5.51 |
| BMD |  |  |  |  |  |  |
| 10mg | 0.48 | 0.19 - 1.21 | 0.7 | 0.14 - 3.41 | 0.98 | 0.35 - 2.71 |
| 5mg | 0.3 | 0.12 - 0.76 | 0.44 | 0.09 - 2.14 | 0.63 | 0.23 - 1.73 |
| BMC |  |  |  |  |  |  |
| 10mg | 1.34 | 0.51 - 3.48 | 0.47 | 0.04 - 4.93 | 1.63 | 0.52 - 5.08 |
| 5mg | 0.88 | 0.32 - 2.39 | 0.88 | 0.19 - 4.16 | 0.87 | 0.23 - 3.25 |
| LTM |  |  |  |  |  |  |
| 10mg | 1.67 | 0.61 - 4.56 | 3.75 | 0.73 - 19.14 | 1.07 | 0.29 - 3.95 |
| 5mg | 1.46 | 0.54 - 3.99 | 1.47 | 0.29 - 7.51 | 1.46 | 0.39 - 5.40 |

However, in conducting these analyses, we observed anecdotally that changes between 24 and 48 weeks were minimal relative to changes from baseline to 24 weeks. This was confirmed by findings of non-significant differences in dosing groups by paired t-tests for the percent change measures of body composition at 24 and 48 weeks, which were again unchanged when separating the analyses by gender (**Supplementary Table S4**). We thus simplified analyses further and performed one-way ANOVA of the change across dosing groups at 24 weeks and 48 weeks. While these remained non-significant for groups overall (24 weeks: VAT: *F(2, 102)* = 0.125, *p* = 0.883, LTM: *F(2, 101)* = 0.877, *p* = 0.419, BMC: *F(2, 98) =* 0.206, *p* = 0.814, BMD: *F(2, 99)* = 0.666, *p* = 0.516; 48 weeks VAT: *F(2, 86)* = 0.335, *p* = 0.716, LTM: *F(2, 87)* = 2.276, *p* = 0.109, BMC: *F(2, 83) =* 1.589, *p* = 0.210, BMD: *F(2, 107)* = 0.365, *p* = 0.695; **Supplementary Table S4**), separating groups by gender in this manner revealed significant differences in LTM (but no other body composition measures) for females across dosing groups after 24 weeks (*F(2, 36)* = 4.208, *p* = 0.023, *ε^2^* = 0.144) and 48 weeks (*F(2, 30)* = 5.052, *p* = 0.013, *ε^2^* = 0.202), with Bonferroni-corrected post-hoc analyses revealing the 10mg group had significant increases in LTM at both timepoints relative to both placebo (24 week: *md* = 3.60472 (95% CI = 0.0913 - 7.1182), *p* = 0.043; 48 week: *md* = 6.194 (95% CI = 0.8773 - 11.5105), *p* = 0.018) and 5mg groups (24 week: *md* = 3.774 (95% CI = 0.3271 - 7.2212), *p* = 0.028; 48 week: *md* = 5.565 (95% CI = 0.5311 - 10.5979), *p* = 0.026; **Table 2**, **Figure 1a, Supplementary Figure S4a**). Interestingly, while with these same methods, males showed differences in VAT at 24 weeks (*F(2, 63)* = 3.548, *p* = 0.035, *ε^2^* = 0.073) with improvements in 5mg relative to 10mg (*md* = −19.520 (95% CI = −37.6513 - −1.3893), *p* = 0.031) but not placebo (*md* = −11.866 (95% CI = −30.4152 - 6.6813), *p* = 0.362), this was not maintained at 48 weeks (*F(2, 54)* = 0.625, *p* = 0.539, *ε^2^* = 0.088; *md* = −13.968 (95% CI = −45.2525 - 17.3168), *p* = 0.825; **Table 2**, **Figure 1b, Supplementary Figure S4b**). While no other measures showed significant differences, trending differences were observed in BMC at 48 weeks in males (*F(2, 52)* = 2.949, *p* = 0.061, *ε^2^* = 0.221), specifically for improvements in 10mg versus 5mg groups (*md* = 2.580 (95% CI = −0.0600 - 5.2198), *p* = 0.057) but not placebo (*md* = 1.383 (95% CI = −1.1092 - 3.8757), *p* = **0.527; Table 2, Figure 1c-d, Supplementary Figure S4c-d**).

**Figure 1.**
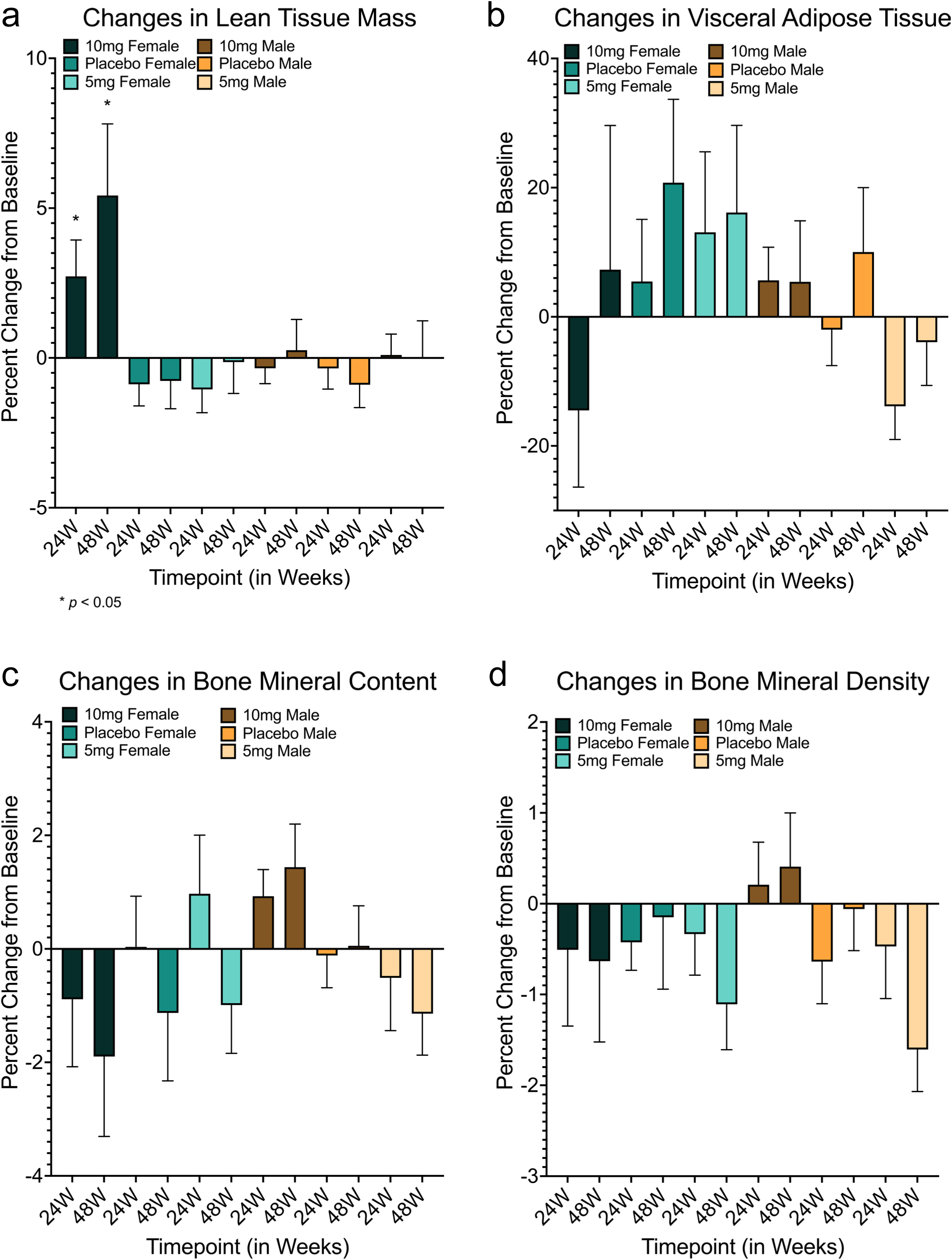
Changes in body composition measures in response to rapamycin use. Females using 10mg of rapamycin had significant improvements in lean tissue mass at 24 and 48 weeks relative to both placebo (24 week: *md* = 3.60472 (95% CI = 0.0913 - 7.1182), *p* = 0.043; 48 week: *md* = 6.194 (95% CI = 0.8773 - 11.5105), *p* = 0.018) and 5mg groups (24 week: *md* = 3.774 (95% CI = 0.3271 - 7.2212), *p* = 0.028; 48 week: *md* = 5.565 (95% CI = 0.5311 - 10.5979), *p* = 0.026) **(a)**. Improvements in visceral adiposity (measured by VAT) were clear for males in the 5mg cohort relative to the 10mg cohort (*md* = −19.520 (95% CI = −37.6513 - −1.3893), *p* = 0.031) but not placebo (*md* = −11.866 (95% CI = −30.4152 - 6.6813), *p* = 0.362) at 24 weeks, but reverted to non-significance after 48 weeks **(b)**. While no other measures showed significant differences **(c, d)**, trending differences were observed in BMC for males at 48 weeks in 10mg versus 5mg groups (*md* = 2.580 (95% CI = −0.0600 - 5.2198), *p* = 0.057) but not placebo (*md* = 1.383 (95% CI = −1.1092 - 3.8757), *p* = 0.527) **(c)**. *md* = mean difference, * = p ≤ 0.05. Error bars represent standard error of the mean.

**Table 2.**
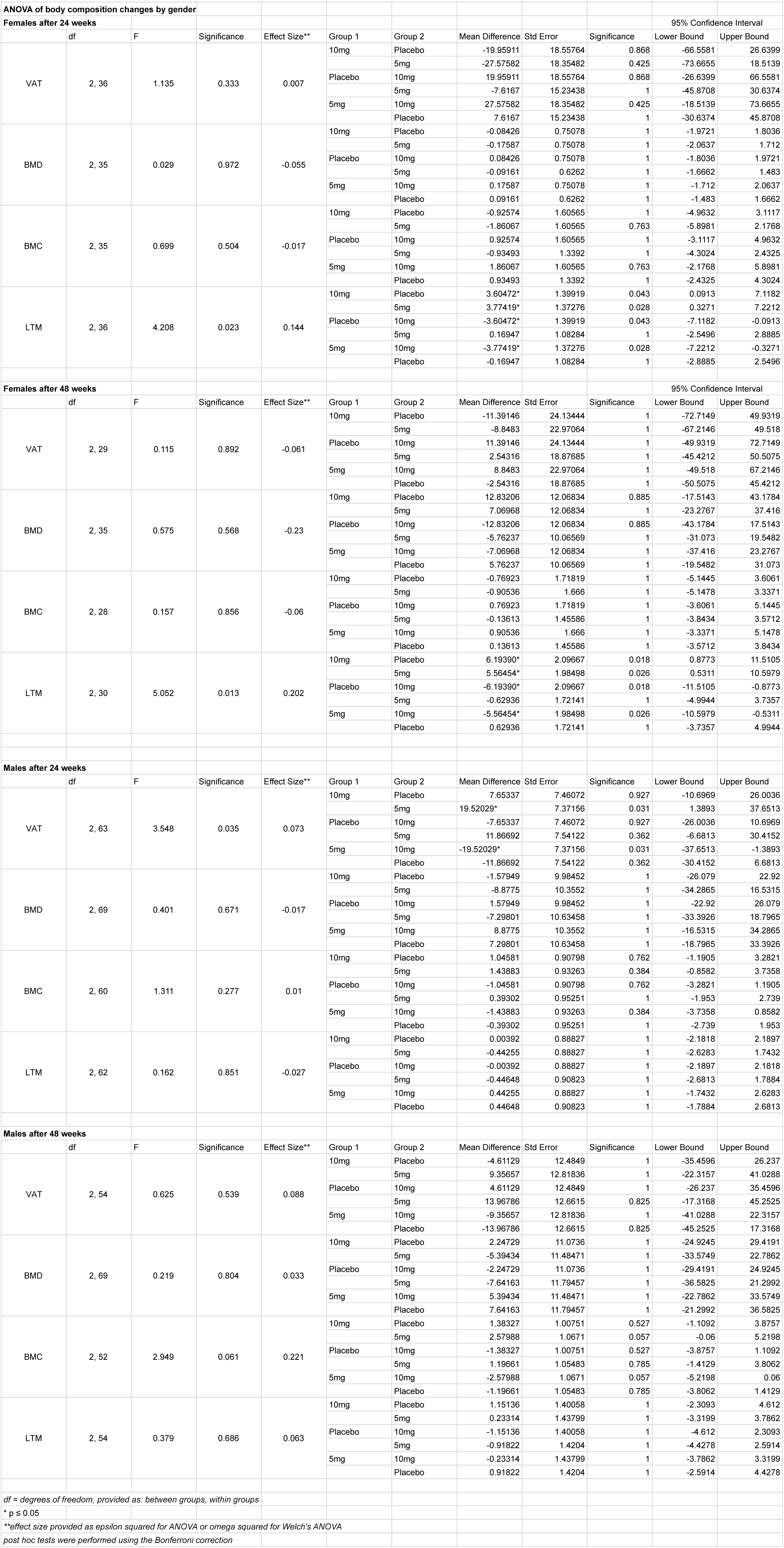
Changes in body composition by DXA scan after 24 and 48 weeks.

Comprehensive blood work panels were completed for all participants at 0 weeks, 24 weeks, and 48 weeks of the study. Repeated measures mixed ANOVA showed no significant changes for most values across time (complete table of results in **Supplementary Table 5**), though consistent patterns of change were observed for some measures. Among these were RBCs (*F(4, 198)* = 2.677, *p* = 0.033, η_p_ ^2^ = 0.051), which Bonferroni corrected post-hoc analyses suggested stemmed from a small (yet still within healthy range) increase in RBC values from 0 to 48 weeks in the 5mg treatment group (*md* = 0.109 (95% CI = −0.189 - 0.003), *p* = 0.042). Separating the groups by gender did not reveal any additional changes for RBCs, however, it did reveal increases (again within healthy range) in BUN levels for males in the 10mg treatment group after 48 weeks (*F(4, 102)* = 2.805, *p* = 0.030, η_p_ ^2^ = 0.099; *md* = 2.222 (95% CI = 0.161 - 4.238), *p* = 0.031).

A number of the initial analyses of blood markers were strongly significant only for changes across time (**Supplementary Table S5**), and changes in values of Hemoglobin A1C, calcium, and CO2 drew particular notice. To more fully explore these effects in the limited context of this study, we again performed repeated measures ANOVA with Bonferroni-corrected post-hoc analyses for each group (**Supplementary Table S6**). While all changes we observed remained within healthy range, Hemoglobin A1C had small increases at 48 weeks in the 5mg cohort only (*F(2, 66)* = 5.630, *p* = 0.006, η_p_ ^2^ = 0.146; *md* = 0.82 (95% CI = 0.015 - 0.150), *p* = 0.013). Separating the groups by gender revealed this was specific to the males in the 5mg cohort (*F(2, 114)* = 4.821, *p* = 0.010, η_p_ ^2^ = 0.078; *md* = 0.059 (95% CI = 0.006 - 0.112), *p* = 0.024). However, no significant changes were observed in glucose or insulin levels (**Supplementary Table S5**). Calcium was seen to change significantly for both 10mg treatment (*F (2, 52)* = 6.909, *p* = 0.002, η_p_ ^2^ = 0.210) and placebo groups (*F (2, 56)* = 4.185, *p* = 0.02, η_p_ ^2^ = 0.130), however, post-hoc analyses suggested that while a transient decrease occurred at 24 weeks (*md* = −0.186 (95% CI = −0.319 - −0.054), *p* = 0.004) and returned to non-significance at 48 weeks for placebo (*md* = −0.100 (95% CI = −0.277 - 0.104), *p* = 0.775), the 10mg group had significant calcium decreases only after 48 weeks (*md* = −0.200 (95% CI = −0.332 - −0.068), *p* = 0.002). Separation by gender revealed this effect to be specific to males (*F(2, 40)* = 3.827, *p* = 0.030, η_p_ ^2^ = 0.161; *md* = −0.167 (95% CI = −0.317 - −0.017), *p* = 0.027). Interestingly, carbon dioxide levels decreased only in the treatment groups (5mg: *F(2, 54)* = 3.891, *p* = 0.026, η_p_ ^2^ = 0.126; 10mg: *F(2, 52)* = 7.492, *p* = 0.001, η_p_ ^2^ = 0.224), with post-hoc tests suggesting the 5mg group experienced changes from 0 to 24 weeks (*md* = −1.250 (95% CI = −2.369 - −0.131), *p* = 0.025) that rose again slightly but non-significantly at 48 weeks (*md* = −0.750 (95% CI = −1.884 - 0.384), *p* = 0.309), while the 10mg group steadily decreased carbon dioxide levels over the course of the study (24 weeks: *md* = −1.185 (95% CI = −2.208 - −0.163), *p* = 0.019; 48 weeks: *md* = −1.222 (95% CI = −2.111 - −0.334), *p* = 0.005)). Separation of effects by gender did not reveal meaningful patterns of gender-specific differences by treatment group (**Supplementary Table S6**).

In the interest of comprehensively evaluating rapamycin responses in our participants, we submitted a subset of samples for epigenetic aging analysis (TruAge from TruDiagnostic, n = 24, 9 female and 15 male) and gut microbiome analysis (Gut Health Test from Thorne, n = 81, 31 female and 50 male) for individuals who consented to additional testing. Within the epigenetic testing results, we saw no meaningful significant changes between groups. In the gut microbiome analyses, no significant differences were observed with repeated mixed measures ANOVA for groups across time, regardless of the separation of groups by gender (**Supplementary Table S7**). Simplified analysis of repeated measures ANOVA for each treatment group also suggested no significant differences in gut microbiome health (**Supplementary Table S7**), though separation by gender suggested small significant increases after 48 weeks in gut dysbiosis in males in the 10mg treatment group (*F (1, 18)* = 4.729, *p* = 0.045, η_p_ ^2^ = 0.228; *md* = 2.235 (95% CI = 0.056 - 4.414), *p* = 0.045), and trends of increased intestinal permeability in females in the 10mg group (*F (1, 4)* = 6.641, *p* = 0.062, η_p_ ^2^ = 0.624; *md* = 3.020 (95% CI = −0.234 - 6.274), *p* = 0.062, **Supplementary Table S7**). However, it should be noted that sample sizes for all groups for epigenetic and gut microbiome health were quite small, and larger sample sizes in future studies would lend more meaningful insight into these measures.

In addition to biological measures of health, validated surveys of self-reported well-being and health (the SF-36 and WOMAC scales) were administered to study participants at 0 weeks, 24 weeks, and 48 weeks. While repeated mixed measures ANOVA was non-significant for changes in treatment groups across time for all WOMAC measures (**Supplementary Table S8**) and SF-36 measures (**Supplementary Table S9**), separating SF-36 scores by gender suggested significant improvements in measures of pain for females over time (*F(4, 66)* = 3.331, *p* = 0.015, η_p_ ^2^ = 0.168, **Table 3**, **Figure 2a**), which Bonferroni-corrected post-hoc analyses revealed was driven by females in the 10mg treatment group at both 24 and 48 weeks (24 weeks: *md* = 6.765 (95% CI = 1.315 - 12.215), *p* = 0.011; 48 weeks: *md* = 8.071 (95% CI = 3.044 - 13.098), *p* < 0.001). Additionally, changes across time were significant for SF-36 measures of Emotional Well-being (*F(2, 184)* = 5.188, *p* = 0.006, η_p_ ^2^ = 0.053) and General Health (Greenhouse-Geisser corrected *F(1.710, 157.342)* = 6.717, *p* = 0.003, η_p_ ^2^ = 0.068). Deeper exploration of these findings with repeated measures ANOVA for each dosing group suggested that these effects stemmed from improvements in the 5mg and placebo groups after 48 weeks for Emotional Well-being (5mg: *F(2, 66)* = 3.987, *p* = 0.023, η_p_ ^2^ = 0.108; *md* = 5.176 (95% CI = 0.056 - 10.297), *p* = 0.047; placebo: *F(2, 58)* = 4.265, *p* = 0.019, η_p_ ^2^ = 0.128; *md* = 4.267 (95% CI = 0.432 - 8.102), *p* = 0.025; **Figure 2b**), however, improvements in General Health reports were specific to the 5mg group (*F(1.757, 57.994)* = 6.582, *p* = 0.004, η_p_ ^2^ = 0.166), and increased at 24 weeks and remained relatively constant thereafter (24 weeks: *md* = 5.882 (95% CI = 0.388 - 11.376), *p* = 0.033; 48 weeks: *md* = 5.882 (95% CI = 1.350 - 10.415), *p* = 0.007; **Figure 2c**). Separation by gender did not impact these findings further (**Table 4, Supplementary Table S9**).

**Figure 2.**
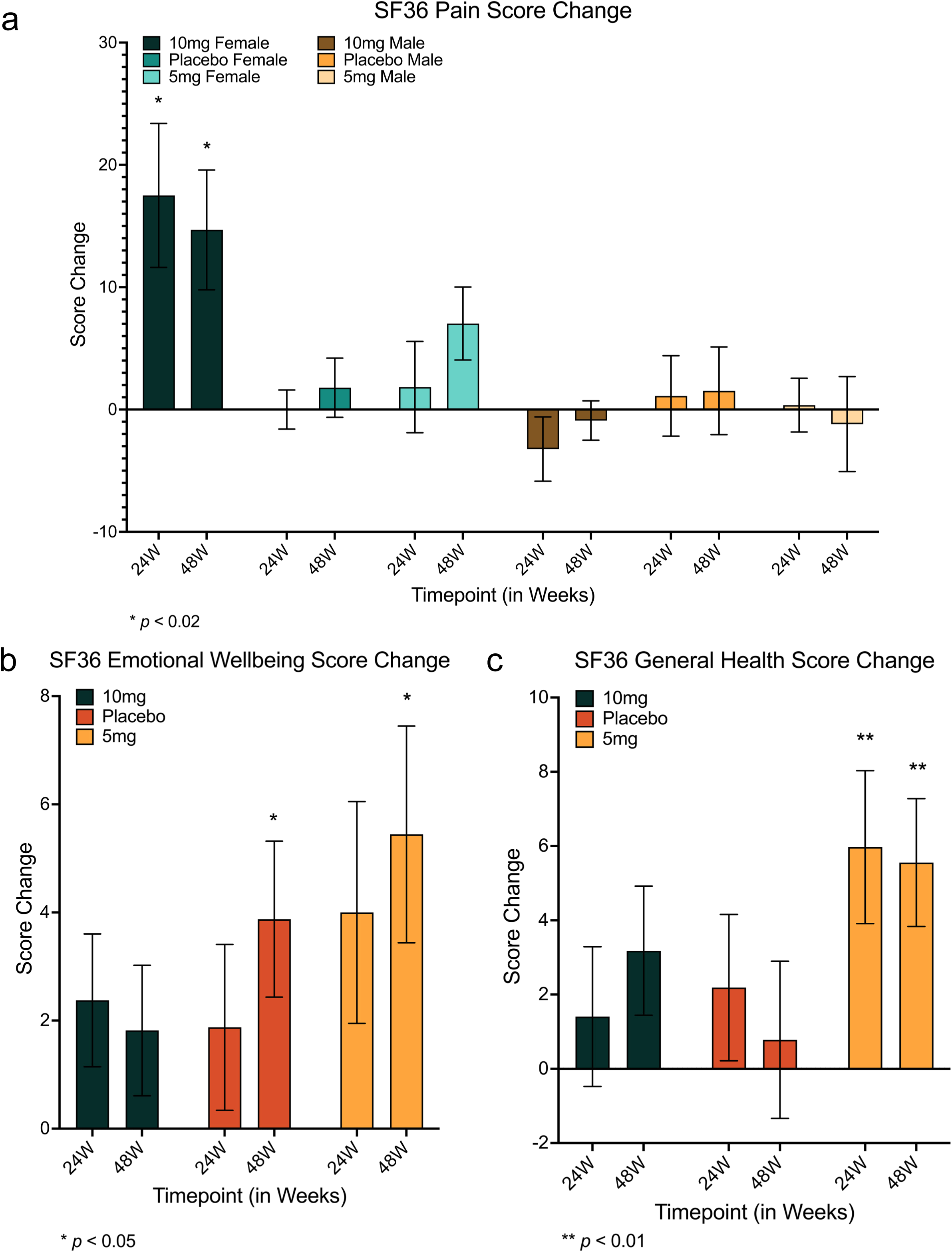
Changes in self-reported survey scores of quality of life and health. Females using 10mg of rapamycin again had significant improvements in self-reported measures of pain at both 24 and 48 weeks (24 weeks: *md* = 6.765 (95% CI = 1.315 - 12.215), *p* = 0.011; 48 weeks: *md* = 8.071 (95% CI = 3.044 - 13.098), *p* < 0.001) **(a)**. Additionally, improvements in measures of Emotional Well-being were seen for 5mg rapamycin users and placebo groups after 48 weeks (5mg: *md* = 5.176 (95% CI = 0.056 - 10.297), *p* = 0.047; placebo: *md* = 4.267 (95% CI = 0.432 - 8.102), *p* = 0.025) **(b)**, however, improvements in General Health reports were specific to the 5mg rapamycin group, increasing at 24 weeks and remaining relatively constant thereafter (24 weeks: *md* = 5.882 (95% CI = 0.388 - 11.376), *p* = 0.033; 48 weeks: *md* = 5.882 (95% CI = 1.350 - 10.415), *p* = 0.007) **(c)**. *md* = mean difference, * = p ≤ 0.05, ** p ≤ 0.01. Error bars represent standard error of the mean.

**Table 3.**
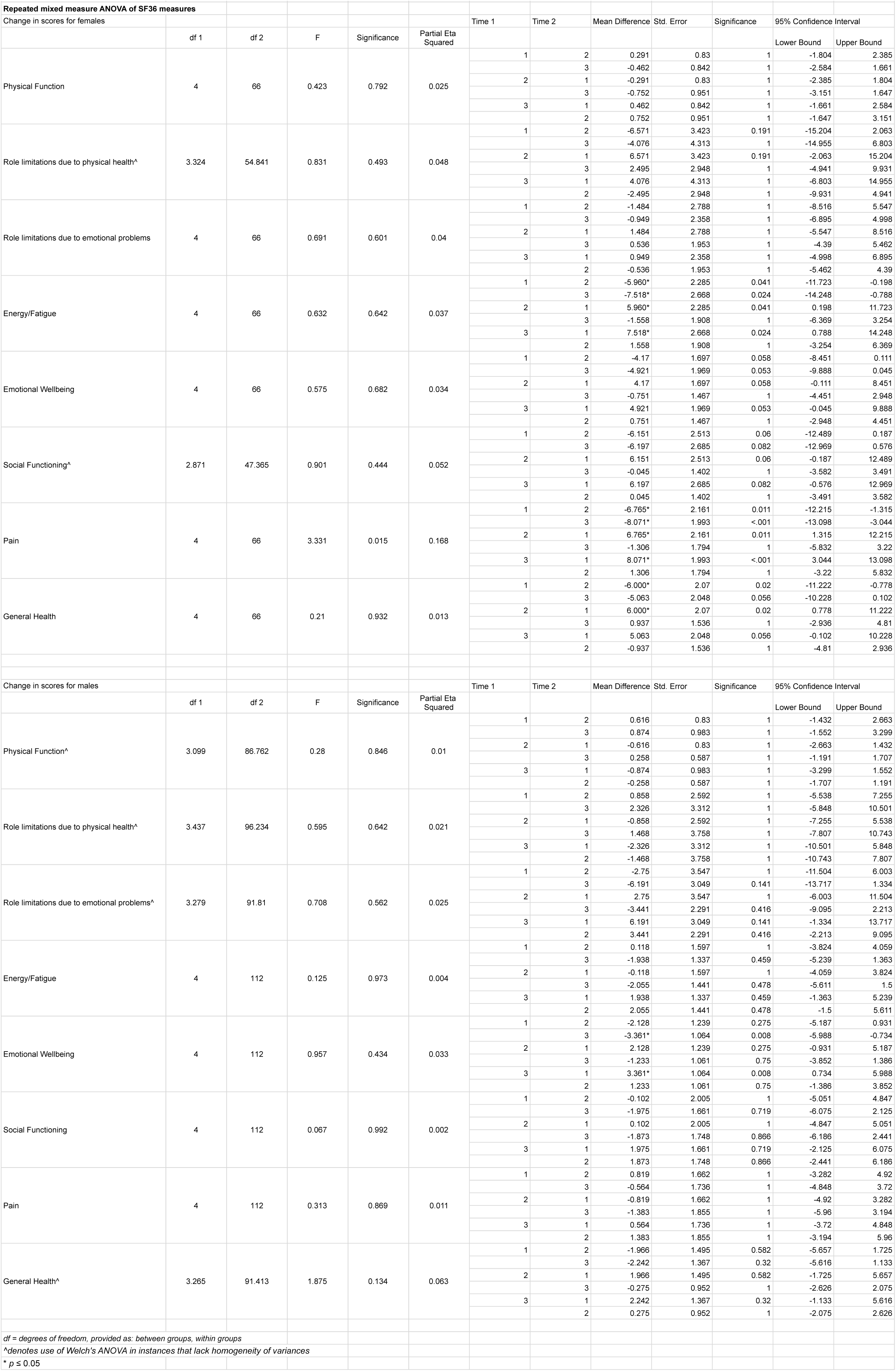
Changes in SF-36 self-reported measures of well-being over 48 weeks.

## Discussion

Few clinical trials to date have evaluated the effects of rapamycin and its derivatives in generally healthy individuals, and those that have been conducted are often challenged by small cohort size, short-term follow-up, or both. While the most robust of these studies have suggested improvements in age-related immune decline in healthy elderly individuals administered low-dose everolimus for 6 to 16 weeks [39], many questions regarding low-dose rapamycin for supporting healthy aging in normative aging individuals remain. The PEARL trial represents one of the largest efforts to date for evaluating the long-term safety and efficacy of low-dose rapamycin for addressing multiple aspects of age-related decline in a normative aging cohort.

The primary goal of the current study was to evaluate the relative safety of low-dose rapamycin use over 48 weeks, and to evaluate whether any clear patterns of concerning side effects emerged in a preliminary cohort. Overall, reports of adverse events (AEs) were relatively consistent across all groups. While rapamycin users appeared to have more GI symptoms than placebo users, no other clear patterns of AEs for rapamycin users emerged. AEs resulting in participant study withdrawal or serious AEs (SAEs) were also similar across groups, with many of the most severe outcomes in the placebo groups. Particular attention was given to immune challenge symptoms for rapamycin users; however, overall reports of cold/flu-like illness and slowed recovery were similar across all groups. There was a single report of anemia in the entire study, in a participant in the 5mg treatment group. While it resolved with treatment and did not recur, we highlight the incidence given interest in this specific outcome for low-dose rapamycin users.

Efficacy was also explored in this study, however, findings are limited by the small relative cohort size, and should be both replicated and extended before definitive guiding conclusions are drawn. Nonetheless, we saw strong improvements in lean tissue mass and self-reported pain symptoms for women taking 10mg of compounded rapamycin (equivalent to ∼2.86 mgs of generic Sirolimus), and promising trends of improvement on other measures of self-reported well-being for both genders (general health and emotional well-being), as well as in bone mineral content measures for males. These effects are largely in keeping with the suggested benefits of low-dose rapamycin use in the longevity community, and provide some measure of clinically validated support for rapamycin’s reputed effects on this front despite the small sample numbers and markedly lower-than-intended dosing. Indeed, these caveats may lend even greater support to the likelihood that rapamycin has longevity benefits, as any evidence of efficacy with such relatively low doses and small numbers is decidedly unexpected. While future studies will be required to more fully understand these effects, and should include a broader dosing range as well as a larger cohort, these findings provide a foundation upon which to build further investigation into the health and longevity effects of low-dose rapamycin.

Interestingly, we observed a broad range of rapamycin responses in evaluated outcome measures for individuals in our study. While our study cohort was particularly health motivated (less likely to drink, smoke, or eat unhealthy diets, and more likely to be active in self-reports) and thus improvements were also seen in the placebo cohort over the study period, we could identify a number of participants using rapamycin on an *n* of 1 scale who had substantial improvements on our evaluated outcome measures. While logic would suggest that this is likely due in part to the influence of varied lifestyle factors (such as diet, alcohol consumption, sleep quality, and activity levels) inherent in a real-world study cohort, the self-reported evaluations of these measures have not correlated with any responses that we have seen to date in any of our studies. However, we are currently investigating a larger cohort of rapamycin users with more objective measures of these factors, in an effort to understand these impacts more fully. In the intervening time, we strongly suggest personalized rapamycin dosing, and continual routine monitoring of blood rapamycin levels in users to ensure maximal benefits, until such time as the optimal longevity dosing dynamics for rapamycin are more clearly understood.

Expanding dosing ranges in future studies will be of particular importance, as while the original design of the current trial intended to utilize a broader range of rapamycin doses, we learned of potential differences in the bioavailability of compounded rapamycin relative to generic from a valued colleague after this trial had already begun. The PEARL trial was temporarily paused, to begin a separate study, with a largely independent cohort of participants, into the bioavailability of different rapamycin formulations (results available on MedRxiv [44]). From this work, we learned that the compounded rapamycin administered for the PEARL study was only equivalent to approximately 28% of the intended dose. While we were sufficiently confident in the bioavailability of the compounded formulation to continue the trial, it was with the understanding that the equivalent dosing was substantially lower than originally intended. Thus, it will be important to explore a broader range of doses to fully understand their impacts in future work.

Taken together, findings from the PEARL trial are the largest and longest to date for evaluating the safety and efficacy of low, intermittent “longevity doses” of rapamycin on healthy aging through the measurement of clinically relevant healthspan metrics. Our findings provide evidence that these rapamycin regimens are well tolerated with minimal adverse effects when administered for at least one year within normative aging individuals. Despite the limited cohort size, we observed some benefits for rapamycin users, particularly women, who had significant improvements in lean muscle mass and self-reported pain. Men had trends of improvement in bone mineral content, however, they did not reach statistical significance. While further investigation into low-dose, intermittent rapamycin’s longevity effects is undoubtedly required and indeed is ongoing, this study provides evidence that rapamycin taken in this manner is relatively safe, and lays the foundation upon which larger and more detailed studies may be developed in the future. Collectively, this and future work aim to collectively build evidence that beyond merely clinical measures of health improvements, rapamycin may promote essential, comprehensive well-being associated with “adding life to years, not just years to life.”

## Supporting information

AEs and SAEs Detail

## Data Availability

All data produced in the present study are available upon reasonable request to the authors

## Acknowledgements

The authors would like to thank the participants who took part in this study. Financial support from our generous donors, particularly Vitalik Buterin, Micah Zoltu, Brad Armstrong, and anonymous gifts, administrative support, and article publishing charges were provided by AgelessRx. Clinical trial oversight was conducted by PHAGE corporation. We extend special thanks to Mikhail Blagosklonny, James Watson, Alan Green and Thorne for their participation and support in conducting this trial.

## Author contributions

Virginia Lee, Andy Nyquist, Anar Isman, and Sajad Zalzala designed and implemented the study. Girish Harinath and Stefanie Morgan performed data analysis. Girish Harinath, Virginia Lee, Andy Nyquist, Mauricio Moel, Stefanie Morgan, Anar Isman, and Sajad Zalzala wrote and edited the manuscript. All work was supervised by Stefanie Morgan, Anar Isman, and Sajad Zalzala. Corresponding author is Stefanie Morgan.

---

**Supplementary Figure S1.**
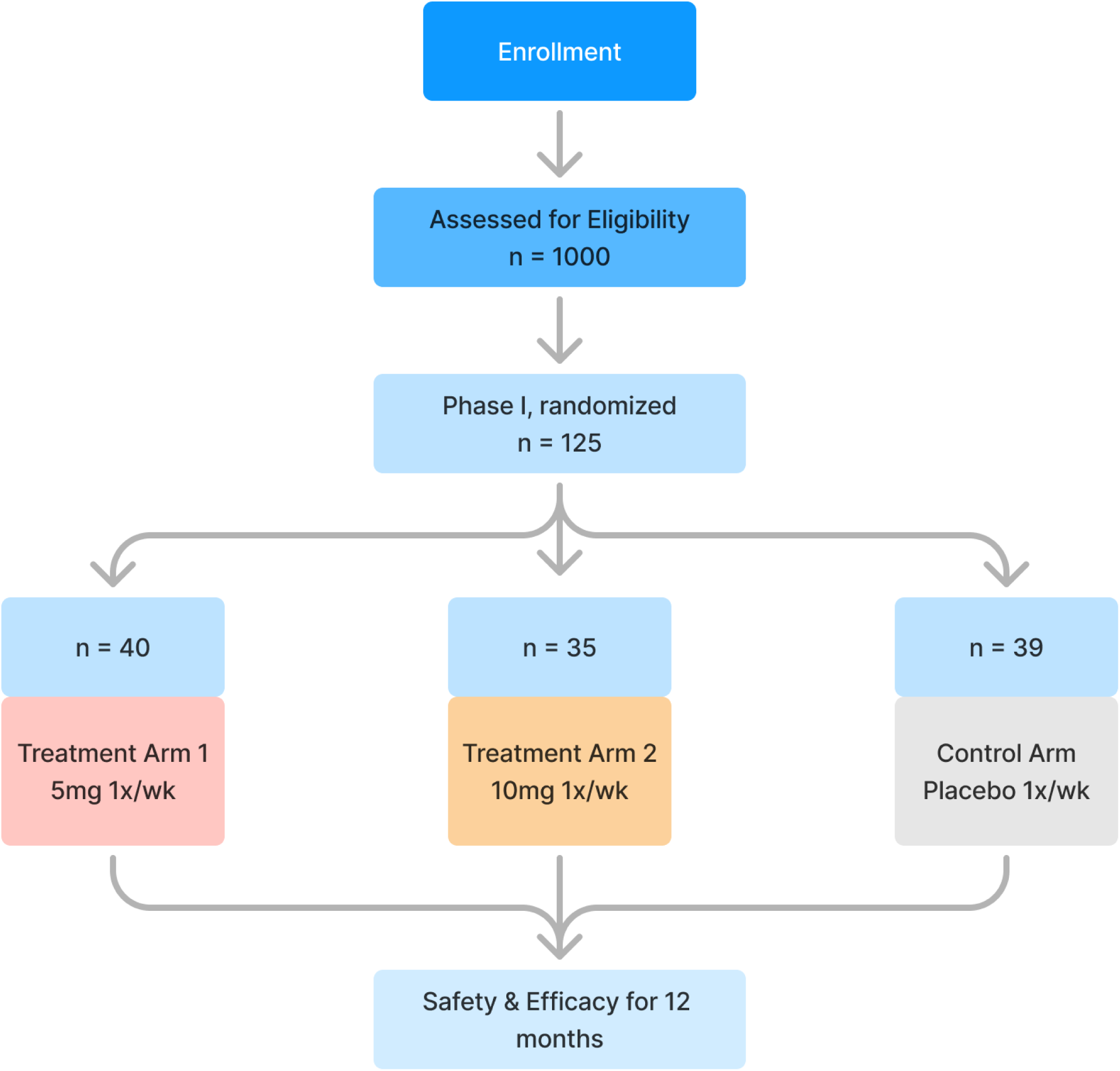
PEARL trial design. Schematic of trial enrollment, participant screening, randomization, and followup (a).

**Supplementary Figure S2.**
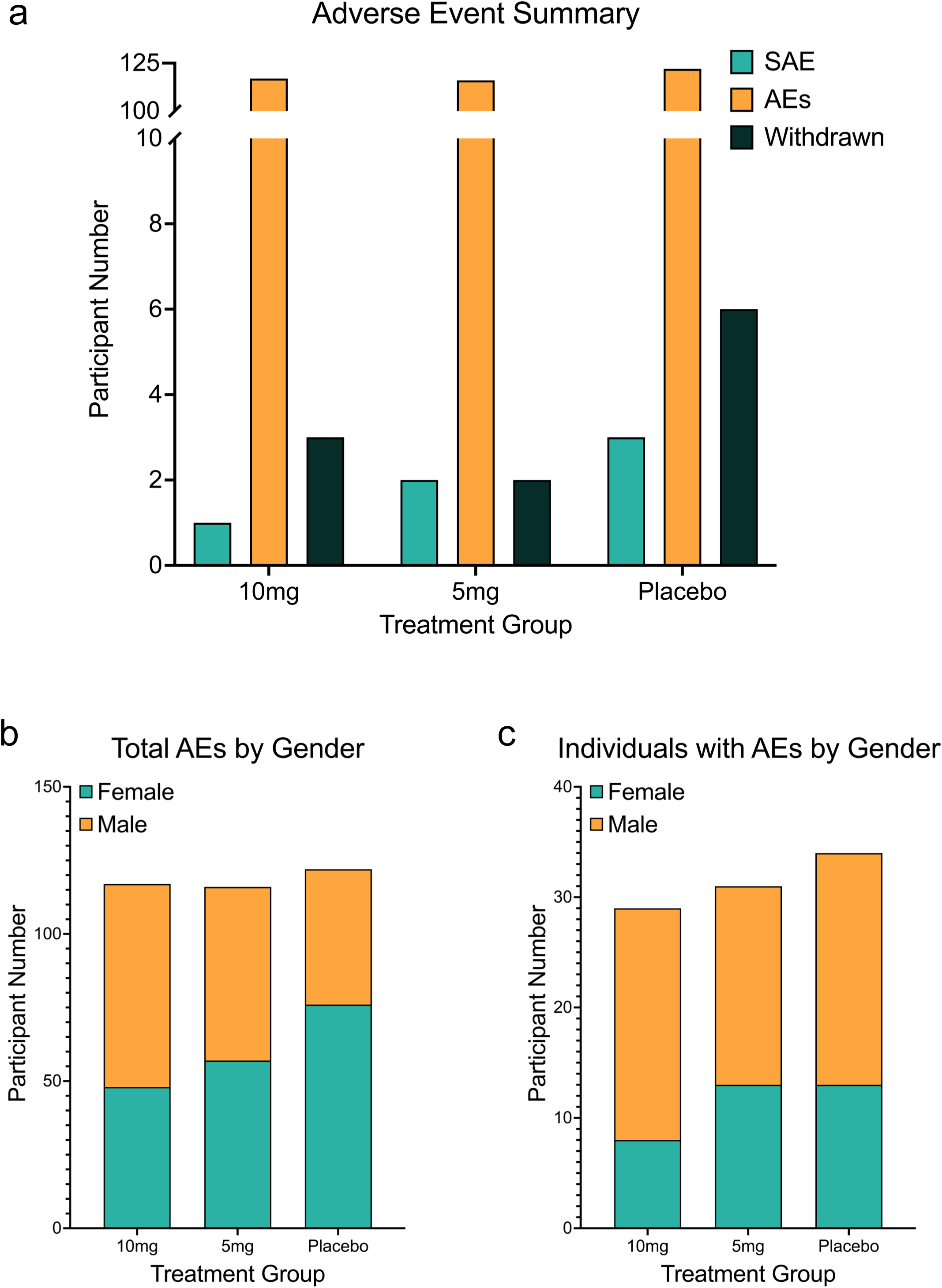
Summary of adverse events and types for PEARL participants. Participants across all groups reported a similar number of incidences of adverse events, with the highest rates of serious adverse events in placebo users **(a)**. Adverse event numbers were similar by gender for all groups **(b)**, and in total number of participants experiencing adverse events in each group **(c)**.

**Supplementary Figure S3.**
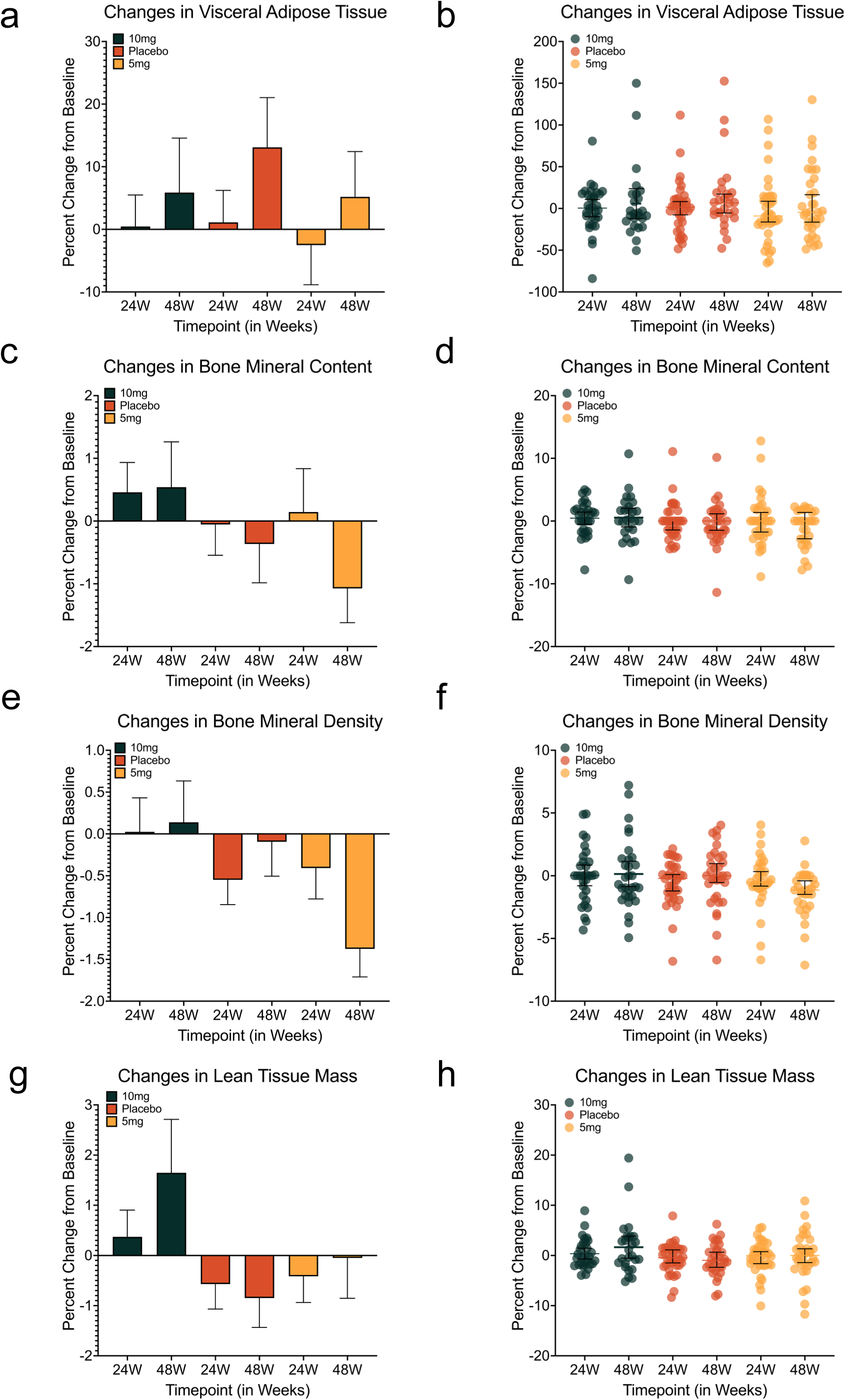
Changes in body composition during the PEARL trial. Body composition measures did not change significantly over the course of the trial for groups as a whole, despite trending differences of improvement for means in some measures **(a-d)**. However, individual changes during the study period were widely varied across all doses and groups **(e-h)**. Error bars represent standard error of the mean.

**Supplementary Figure S4.**
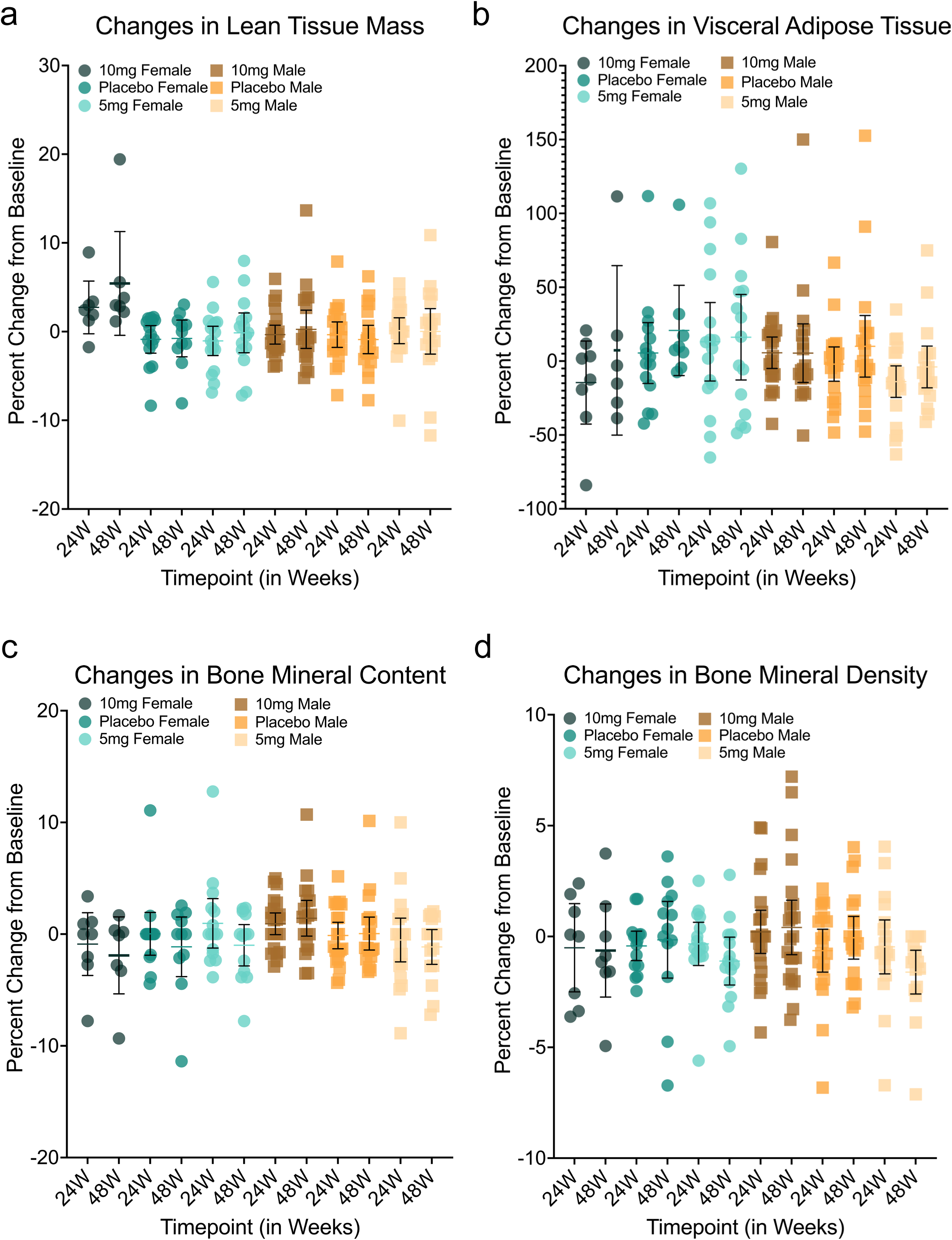
Heterogeneity of individual response in body composition for each gender during the PEARL trial. Individual responses for measures of body composition change span a range of values for each dose and gender (a-d). Bars represent the 95% CI, dots represent individuals.

