## Supplementary material for "Safety and efficacy of rapamycin on healthspan metrics after one year: PEARL Trial Results": AEs and SAEs Detail

**Supplementary File 1: AE and SAE Report Detail**

**Withdrawn:**

- 10mg
    - The participant experienced a sore throat and low-grade fever (~100°F) approximately 24 hours after their first dose, which improved after a few days. They had a prior respiratory infection before the dose. They also reported one small acne spot and a canker sore that resolved quickly. Further survey responses indicated new acne, a canker sore, and gastrointestinal (GI) issues, specifically a "burning sensation in the stomach and nausea," along with a sore throat, cough, and earache. These symptoms typically began a few days after taking the capsule and lasted 2-3 days.
    - A pharmacy error resulted in the participant receiving a full dose instead of the intended half dose. After meeting with clinical trial associates, the correct half dose was sent for future administration, but only after the participant's symptoms resolved. For statistical analysis, the following events will be documented separately: general cold/flu with GI symptoms, acne, canker sore, and a second GI event.
    - During a routine check-in, the participant reported the return of GI symptoms (burning sensation) 3-4 weeks after taking the half dose. They decided to discontinue the treatment and withdraw from the trial.
- Placebo
    - The participant reported gastrointestinal (GI) symptoms including loose stools, increased flatulence, up to three bowel movements per day, vague central abdominal pain, malaise, and lack of energy. They also noted a decline in hip health (pain and mobility issues), which had previously improved, and an increase in blood pressure (from 113/78 to 130/85). There were no fever or other symptoms, but these issues affected their ability to work and exercise. The participant had a history of gastroenteritis and ulcers, and a clear colonoscopy from December 2021. They attempted a 36-hour fast and a short course of over-the-counter proton pump inhibitors (PPIs), neither of which resolved the symptoms. They also engaged in intensive physical therapy for the hip problems but did not seek a provider for further diagnostics.
    - After speaking with trial staff, the participant agreed to discontinue Rapamycin. Follow-up surveys indicated steady improvement in their GI symptoms, with resolution. However, the participant expressed reluctance to restart the medication and requested to withdraw from the trial.
- Placebo
    - Participant reported “Cough, Mild pains, Headache” via survey. Follow-up reported COVID-19 positive result. Staff requested participant follow CDC guidelines and delay 4-week Quest panel. Next survey reported: “Congestion, Cough, Fever, GI, Mild pains, Respiratory, Headache, Joints, Tightness in

breath, Felt I couldn't digest". Follow-up confirms no symptoms for 06/2022 Quest panel.
    - Participant reported "Skin, Blistering/Irritation/ Itching/Reddening" via survey. Participant reported bruising more than usual without memorable cause and a rash/cut on the hand not healing as the participant expects. Upon follow-up, Participant requested to withdraw and was asked to schedule a debriefing appointment.
    - 06/2022 follow-up reports "eye pain" like "sand in the eye" with painful blinking. Potentially if the rash on hand was infectious, it could have infected eye. Participant has not sought medical attention from a PHP, dermatologist, ophthalmologist, or urgent care. However, all symptoms have resolved.
- 10mg
    - Participant reported they will need to have a small bladder stone removed in the future. Participant did not report any symptoms when staff asked to describe them. Participant confirmed that their urologist saw the stone during a BPH check-up and advised removal.
    - Participant expressed desire to withdraw due to assumption of being on placebo.
- Placebo
    - Participant noted increase in balance issues during Week 2 check-in.
    - Participant noted they have a pre-existing condition being treated that results in "balance issues". No medical conditions were noted in the PHP Screening documentation. Participant was asked to note when the issue changes in frequency/severity.
    - Participant reports Neurological/Behavioral issues, Dizziness, Slurred speech, Faintness/Lightheadedness via survey. Symptoms appear to be a flare of the condition previously mentioned.
    - Participant withdrawing as of 01/2023 due to desire to be sure they are not on placebo.
- 5mg
    - No AE just wanted to withdraw with spouse
- Placebo
    - Participant reports tinnitus via survey. Participant follow confirms tinnitus and hearing loss Dx prior to starting the trial. No treatments or lifestyle changes noted. Participant noted symptoms on survey due to "resurfacing" of symptoms. Participant notified of possibility of permanent hearing loss due to Rapamycin. Participant advised to keep a close eye on symptoms and follow up with audiologist regularly to prevent permanent issues.
    - Participant reports severe muscle and bone pain. Follow-up confirms the issue began 3 years ago, but has been worsening. Participant underwent MRI and was Dx with avascular necrosis, and will require hip replacement surgery.
    - Participant decided to withdraw due to new Dx and Tx plan. Staff supports decision for best outcomes.
- 10mg

    - Participant reports an “unnamed pathogen…, abnormal DNA and several markers for inflammation” was found in their Baseline Thorne gut health kit. Participant advised to seek treatment from their primary care and report any new prescriptions.
    - No symptoms were reported. Therefore, no adverse event has occurred.
    - Participant reports post-COVID-19 symptoms via email. Follow-up reports no positive COVID-19 test and did not provide possible date of exposure, as requested. Unknown if acute symptoms occur before or after first dose, due to lack of response. Participant confirms a “very high” C-reactive protein level, which increases with flu, COVID-19, and most any viral infection.
    - Participant lists current symptoms as: fatigue, post-exercise fatigue, brain fog, and ringing in the ears. The symptoms are rated 6-9/10 bothersome as of 09/2022. Participant rests in bed 16hr/day and has discontinued usual exercise routine. Provider believes symptoms due to long COVID-19.
    - Trial clinician approves temporary discontinue of trial therapeutic and reassessment before continuing in the trial.
    - Temporary dose delay for 2 weeks offered, but participant unresponsive to follow-up emails. Voicemail left with contact info. See communications log.
    - Follow-up response on 11/2022: participant requested to withdraw and was asked to schedule a debriefing appointment.
- 5mg
    - Participant reports “GI” on survey. Follow-up confirms participant is experiencing constipation. Hx of recurrent constipation, but participant notes this case is more severe than before the trial (no bothersome scale given).
    - Participant reports seeking care from PHP and Gastroenterologist, both of whom recommended OTC Miralax. Participant Tx’d with Miralax for 2-3 weeks and symptoms did not resolve. Participant notes not having a bowel movement for up to 1 week during this time.
    - Participant reports being advised to discontinue the trial therapeutic. Follow-ups sent to confirm who recommended this and the dates at which this occurred, as it is not documented.
    - Follow-up confirms their provider believes the issue is the trial therapeutic and advised them to discontinue. After 2 weeks of unreported delayed doses, symptoms improved. The participant decided to withdraw from the trial, under advisement of the provider.
- Placebo
    - Participant reports via email, lumps in their groin. Participant has a personal and family Hx of lipoma. CT performed, inflamed lymph nodes suspected, but not diagnosed.
    - Follow-up denies Hx of inflamed lymph nodes. Lumps are bothersome (did not provide a rating) and hard with no discoloration. There are 3x in total as of 08/24, but began with only 1x ~2-3mo ago. The last one appeared in the past 2 weeks. Participant notes previously unreported hip pain that they attribute as the lumps compressing the groin area.

- Biopsy on 08/24, results expected 08/29. Provider does not believe it is related to Rapamycin. Participant expressed desire to continue in the trial as long as it is approved by trial clinician to do so.
- In 09/2022 follow up, reports Dx of "metastatic, small-cell neuroendocrine adenocarcinoma". Participant advised to discontinue immediately due to upcoming chemotherapy treatments.

- 05/2023 email follow-up with clinical staff, participant reports no cancer from scans and results. Still withdrew from study at this time.

**SAE:**

- 5mg, completed trial.
  - Participant reports 2 days "inpatient" (unable to confirm inpatient status versus admitted for observation) for "anemia" and given "one unit" of blood via transfusion. Respiratory symptoms (i.e. shortness of breath) appear to be due to anemia and not pneumonia.
  - Anemia resolved via blood transfusion and participant sent home from hospital. No further AE reported.
- 5mg, completed trial.
  - Participant reported "Congestion, Cough, Mild pains, Joints, Back/Ribs/Arms/Legs, Mouth irritation"via survey. Follow-ups indicated that illness progressed and participant directed to consult Primary Healthcare Provider (PHP), who prescribed: 4x days of Prednisone, 10x days of Doxycycline hyclate, & Albuterol inhaler BID. Illness was originally documented as an expected adverse event and then upgraded to severe on 03/2022 due to the participant seeking treatment from the provider, and the worsening of symptoms over time rather than improvement. Participant reports this illness was unusually severe compared to previous colds/illnesses. Participant was asked to skip doses while administering prescription medication from Primary Healthcare Provider. Participant skipped one dose, which was logged in the Protocol Deviation Log.
  - Participant took medications as prescribed and reported complete resolution.
- SAE, Placebo, deceased and did not complete study.
  - Participant reported expected adverse event consisting of "GI" via survey (which are exported and reviewed for the whole trial 3-4x/mo). Participant followed up with staff via email detailing flu-like symptoms and rib pain (also expected), which appeared to be resolving at the time of follow-up. Participant did not respond to further emails requesting more information regarding details of symptoms, any treatments taken, and complete resolution. Participant stopped filling out weekly adverse event surveys. It is not uncommon for PEARL participants to feel overwhelmed by emails or be generally technologically avoidant due age, so some leniency is provided in our standard procedures before withdrawing the participant.
  - The PHP later contacted trial staff to confirm hospital records document cardiogenic shock due to myocardial infarction as the cause of death. While

Rapamycin/Sirolimus may cause changes in heart rate, other cardiac events are not reported as a possible side effect. Participant was out of town during this time and did not receive care from local hospital or clinic, which may have caused delay in reporting to trial staff.

- SAE, Placebo, completed trial.
    - Participant reports going to the ER for “a stomach virus”, which presented with nausea , vomiting, and diarrhea. The ER diagnosed the participant with a “small urinary infection” and prescribed Nitrofurantoin mono/mac 100mg caps for 5 days and Ondansetron ODT 4mg tablets. The participant did not require in-patient hospitalization, but felt uncomfortable enough to go to the ER. Symptoms resolved with treatment.
- SAE, 10mg, completed trial.
    - Participant’s spouse reports going to the ER for “a 10/10 sore throat” 07/2023 during follow-up concerning resolution of symptoms previously reported in routine health survey: Cough, tightness of breath, and “Amoxicillin for Strep as of 7-23”. Tightness of breath denied in follow-up with spouse. Spouse also denies fever. Participant treated with OTC cough medicine and Vitamin C prior to going to the emergency room.
    - Participant did not require in-patient hospitalization, but felt uncomfortable enough to go to the ER.
    - ER tested for COVID-19 (negative) and 2x Streptococcus (one “quick test” that was not positive and one culture). There has been no follow-up on the culture as of 07/2023. ER “did not see white spots on [their] tonsils” and there was no fever. ER did not diagnose Strep throat and did not prescribe medications. Participant called PHP and PHP prescribed both Amoxicillin and “pain medication”. Pain medication was out of stock, but participant’s spouse reports symptom improvement 12hrs after Amoxicillin administration.
- AE, Placebo, completed trial.
    - *Note: Participant was not hospitalized and did not require an invasive procedure nor was disabled. Designated not SAE upon 07/14/2024 audit.
    - Via email, participant reported worsening of injury that occurred prior to enrollment. Participant did not report injury before this time, as it was mild pain in right gluteus and assumed to be related to long-distance running. Participant reports an appointment with a provider specializing in Sport Medicine, who diagnosed “Type III-A tear of my semimembranosus hamstring at the ischial tuberosity” via MRI.
    - Participant is receiving treatment via platelet-rich plasma injection and physical therapy. If not fully healed, care plan includes second platelet-rich plasma injection and continued physical therapy. Surgery is an option; however, the participant reported they will not consider it until the other treatments have failed, which will be “for at least 9 months”.

    - Participant advised to be sure to notify care team of possible Rapmycin administration, which may slow healing. If care team decides for the participant to discontinue administration, withdrawal and/or delay of dosing options will be discussed at that time.
